# Community carriage of ESBL-producing *Escherichia coli* and *Klebsiella pneumoniae*: A cross-sectional study of risk factors and comparative genomics of carriage and clinical isolates

**DOI:** 10.1101/2022.11.09.22282110

**Authors:** Niclas Raffelsberger, Dorota Julia Buczek, Kristian Svendsen, Lars Småbrekke, Anna Kaarina Pöntinen, Iren H. Löhr, Lotte Leonore Eivindsdatter Andreassen, Gunnar Skov Simonsen, Norwegian E. coli ESBL Study Group, Arnfinn Sundsfjord, Kirsten Gravningen, Ørjan Samuelsen

## Abstract

The global prevalence of infections caused by ESBL-producing Enterobacterales (ESBL-E) is increasing and for *Escherichia coli* observations indicate that this is partly driven by community-onset cases. The ESBL-E population structure in the community is scarcely described and data on risk factors for carriage are conflicting. Here, we report the prevalence and population structure of fecal ESBL-producing *E. coli* and *Klebsiella pneumoniae* (ESBL-Ec/Kp) in a general adult population, examine risk factors, and compare carriage isolates with contemporary clinical isolates.

Fecal samples obtained from 4999 participants (54% women) ≥40 years in the seventh survey of the population-based Tromsø Study, Norway (2015-2016) were screened for ESBL-Ec/Kp. In addition, we included 118 ESBL-Ec clinical isolates from the Norwegian surveillance program in 2014. All isolates were whole-genome sequenced. Risk factors associated with carriage were analyzed using multivariable logistic regression.

ESBL-Ec gastrointestinal carriage prevalence was 3.3% (95% CI 2.8-3.9%, no sex difference) and 0.08% (0.02-0.20%) for ESBL-Kp. For ESBL-Ec, travel to Asia was the only independent risk factor (AOR 3.47, 95% CI 2.18-5.51). *E. coli* ST131 was most prevalent in both collections. However, the ST131 proportion was significantly lower in carriage (24%) vs. clinical isolates (58%, p<0.001). Carriage isolates were genetically more diverse with a higher proportion of phylogroup A (26% vs. 5%, p<0.001), indicating that ESBL gene acquisition occurs in a variety of *E. coli* lineages colonizing the gut. STs commonly related to extra-intestinal infections were more frequent in clinical isolates also carrying a higher prevalence of antimicrobial resistance, which could indicate clone associated pathogenicity.

**Importance:** ESBL-producing *E. coli* (ESBL-Ec) and *K. pneumoniae* (ESBL-Kp) are major pathogens in the global burden of antimicrobial resistance. However, there is a gap in knowledge concerning the bacterial population structure of human ESBL-Ec/Kp carriage isolates in the community. We have examined ESBL-Ec/Kp isolates from a population-based study and compared these to contemporary clinical isolates. The large genetic diversity of carriage isolates indicates frequent ESBL gene acquisition, while those causing invasive infections are more clone dependent and associated with a higher prevalence of antibiotic resistance. The knowledge of factors associated with ESBL carriage helps to identify patients at risk to combat the spread of resistant bacteria within the healthcare system. Particularly, previous travel to Asia stands out as a major risk factor for carriage and should be considered in selecting empirical antibiotic treatment in critically ill patients.

## Introduction

Extended spectrum β-lactamase (ESBL) producing *Escherichia coli* (ESBL-Ec) and *Klebsiella pneumoniae* (ESBL-Kp) are major contributors to the global burden of disease due to antibiotic resistant bacteria.^1^ The prevalence of infections caused by ESBL-Ec or ESBL-Kp is increasing, even in countries with low antibiotic use and availability on prescription only, like Norway.^2^ The mortality of invasive infections with ESBL-producing Enterobacterales (ESBL-E) ranges from 10-35%, depending on bacterial species, host factors, severity of disease and initial antibiotic therapy.^3,4^ Consequently, ESBL-Ec/Kp are high-priority pathogens when developing new drugs to combat the threat from antibiotic resistant bacteria.^5^

ESBL-Ec/Kp colonization precedes invasive infections^6–8^ and recent reviews report an increasing global prevalence of human ESBL-E carriage in the community, with an 8-fold rise over the past two decades.^9,10^ The highest prevalence is detected in Asian and African regions (20-70%), the lowest in Europe and the Americas (<10%).^9,10^ A study from the USA shows that the increased incidence of ESBL-Ec infections was driven by an increase in community-onset cases^11^, and human-to-human transmission was the attributable source to 60% of community-acquired gastrointestinal ESBL-Ec carriage in the Netherlands ^12^.

Several risk factors for ESBL-E gastrointestinal carriage have been described. However, these have, with a few exceptions^13,14^, mainly been investigated in small and/or selected study populations, such as international travelers^15^, patients with gastroenteritis^16^, patients recruited by general practitioners^17^, persons with recent healthcare contact^18^, pregnant women^19^, children^20^ or persons living in a livestock-dense area^21^. Many studies have identified international travel as a risk factor for ESBL acquisition, whilst the significance of sex, age, antibiotic or proton pump inhibitor use, hospitalization and diet are conflicting.^13–15,17,18,22–24^

We have comprehensive knowledge of the prevalence and population structure of clinical ESBL-E isolates showing a dominance of CTX-M-group ESBL enzymes and the association with specific extraintestinal pathogenic *E. coli* (ExPEC) and multidrug-resistant *K. pneumoniae* high-risk clones.^25–28^ Large-scale genomic studies have identified specific subclades of *E. coli* sequence type (ST) 131 and *K. pneumoniae* ST307 as major contributors to the increasing prevalence of ESBL infections.^25,27,29,30^ Community-based studies from the Netherlands and Sweden also observed a predominance of ST131, but a high genetic diversity within the ESBL-Ec population.^13,14^

Improved knowledge of risk factors for ESBL-E carriage and their population structure in the general human population may provide information for risk stratification and targeted infection control measures. It is important to consider ESBL-E carriage in critically ill septic patients with respect to the choice of empirical antibiotic treatment. The aims of this study were to examine the prevalence of, and risk factors associated with, ESBL-Ec/Kp gastrointestinal carriage in a general adult population in Norway, and to compare the ESBL-Ec population structure with a national collection of clinical isolates.

## Results

We detected gastrointestinal carriage of putative ESBL-Ec/Kp in 188 of 4,999 randomly selected participants who provided fecal samples in the seventh survey of the population-based Tromsø study (Tromsø7)^31^ (Suppl. Table 1, Figure 1). Overall, 87% of participants receiving a sampling kit returned a fecal sample. In total, 180 Ec and nine Kp putative ESBL-positive isolates were isolated from the 188 ESBL screening-positive fecal samples. Both ESBL-Ec and -Kp were detected in one sample.

**Figure 1.**
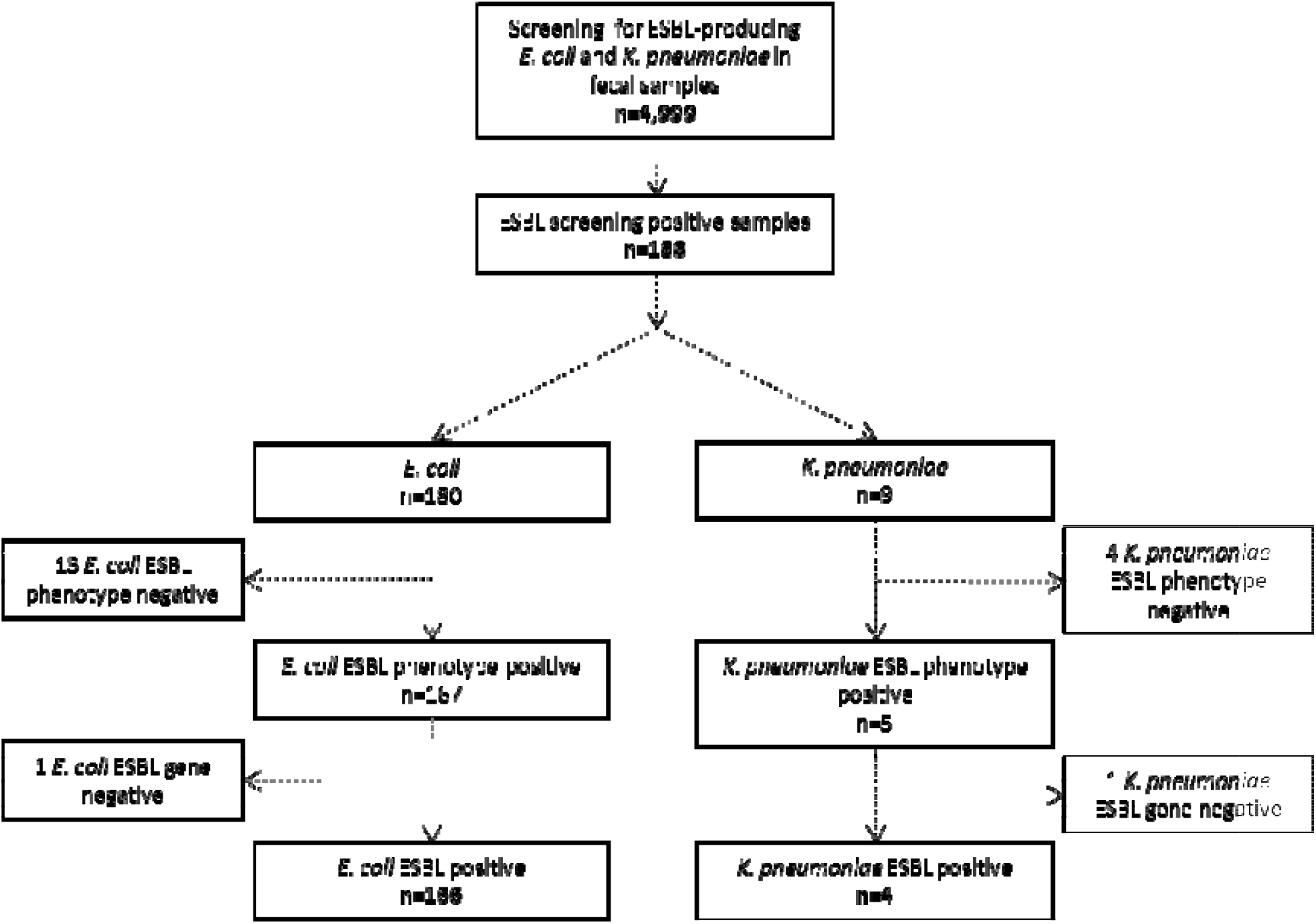
Flowchart and results of fecal sample screening for gastrointestinal carriage of ESBL-producing *E. coli* and *K. pneumoniae* in 4,999 participants in Tromsø7, 2015-2016.

Pheno- and genotypic analyses showed that 14 putative ESBL-Ec isolates were either pheno- and/or genotypically ESBL-negative. Of these, one isolate harbored plasmid-mediated AmpC (*bla*CMY-2). After exclusion of the ESBL-negative isolates, prevalence of ESBL-Ec gastrointestinal carriage was 3.3% (95% confidence interval (CI) 2.8-3.9%, 166 of 4,999 participants); 3.1% (2.5-3.9%) in women and 3.5% (2.8-4.4%) in men.

Among the nine screening ESBL-positive Kp isolates, one harbored *bla*CMY-2 and four did not express an ESBL phenotype. Consequently, four samples were considered positive for ESBL-producing Kp, corresponding to a prevalence of 0.08% (0.02-0.20%), all among male participants.

### Factors associated with gastrointestinal carriage of ESBL-E. coli

We analyzed data from 4,996 participants, excluding those three who were positive for ESBL-Kp only (Suppl. Table 1). Median age was 65 years (interquartile range 58-70 years, no sex difference). In multivariable logistic regression analyses adjusted for all explanatory variables, only travel to Asia the past 12 months was associated with ESBL-Ec gastrointestinal carriage with an adjusted odds ratio (AOR) of 3.47 (2.18-5.51) (Table 1). Among participants reporting hospitalization in the past year, or recent use of antibiotics or acid suppressive medication, we observed a non-significant increase in prevalence of ESBL-Ec.

**Table 1.** ESBL-producing *E. coli* gastrointestinal carriage and associated factors among 4,996 participants in Tromsø7.

| Characteristics | %<br>(ESBL-<br>*E. coli*) | n<br>(ESBL-<br>*E. coli*) | N | AOR | 95% CI | p-value |
| --- | --- | --- | --- | --- | --- | --- |
| Age (years) |  |  |  |  |  | 0.236 |
| 40-49 | 3.6 | 22 | 605 | 1.00 |  |  |
| 50-59 | 4.1 | 33 | 814 | 1.15 | 0.66-2.01 |  |
| 60-69 | 3.2 | 68 | 2,127 | 0.81 | 0.49-1.33 |  |
| 70-84 | 3.0 | 43 | 1,450 | 0.73 | 0.42-1.26 |  |
| Hospitalization past 12 months |  |  |  |  |  | 0.128 |
| No | 3.3 | 142 | 4,341 | 1.00 |  |  |
| Yes | 4.0 | 24 | 593 | 1.43 | 0.90-2.27 |  |
| Antibiotic use past 14 days<sup>a</sup> |  |  |  |  |  | 0.258 |
| No | 3.3 | 158 | 4,831 | 1.00 |  |  |
| Yes | 5.2 | 8 | 153 | 1.58 | 0.72-3.47 |  |
| Acid suppressive medication past 4 weeks |  |  |  |  |  | 0.951 |
| Not used | 3.3 | 123 | 3,760 | 1.00 |  |  |
| ≤weekly | 3.2 | 13 | 402 | 1.02 | 0.57-1.83 |  |
| Every week, but not daily | 3.5 | 9 | 260 | 1.10 | 0.55-2.20 |  |
| Daily | 3.7 | 11 | 297 | 1.20 | 0.63-2.26 |  |
| Travel abroad past 12 months<sup>b</sup> |  |  |  |  |  | <0.001 |
| No | 2.5 | 53 | 2,145 | 1.00 |  |  |
| Other regions (excl. Asia) | 3.2 | 70 | 2,212 | 1.36 | 0.93-1.98 |  |
| Asia exclusively or Asia + other regions | 8.2 | 38 | 461 | 3.47 | 2.18-5.51 |  |
| Traveler's diarrhea past 12 months<sup>c</sup> |  |  |  |  |  | 0.972 |
| No | 3.3 | 156 | 4,721 | 1.00 |  |  |
| Yes | 4.8 | 8 | 166 | 1.01 | 0.48-2.16 |  |

ESBL, extended spectrum β-lactamase; N, denominator; AOR, adjusted odds ratio; CI, confidence interval; AOR adjusted for age, hospitalization past 12 months, antibiotic use past 14 days, acid suppressive medication past 4 weeks, travel abroad past 12 months and traveler's diarrhea past 12 months.
The multivariable model includes 4,488 participants with complete information on all variables.
<sup>a</sup> Have you taken any antibiotics (tablets or oral suspensions, nasal ointments, eye drops or eye ointment) during the past 14 days?
<sup>b</sup> Traveled outside the Nordic countries >1 week duration in the past 12 months.
<sup>c</sup> For each travel abroad the past 12 months, the participants were asked if they had experienced diarrhea in connection with the travel.

Both in the literature and in our previous study^32^, we identified several factors associated with Kp gastrointestinal carriage, overlapping with those associated with ESBL-Ec carriage.^15,22,33,34^ Thus, in 2,972 participants (of the total 4,996 participants in this current study) previously screened for Kp, we performed a multivariable logistic regression analysis including Kp carriage as an additional explanatory variable (Suppl. Table 2). Prevalence of ESBL-Ec gastrointestinal carriage was 2.7% among participants without Kp carriage and 4.8% (p=0.016) among those with Kp carriage. In the full model, AOR was 1.66 (0.99-2.79, p=0.055), indicating a possible association between Kp and ESBL-Ec carriage.

### Comparative analysis of ESBL-E. coli carriage and clinical isolates

To explore the population structure and genomic characteristics of ESBL-Ec in community carriage we whole-genome sequenced (WGS) all 166 isolates. Furthermore, we sequenced a contemporary national collection of 118 clinical ESBL-Ec isolates (NORM 2014) for comparative analysis (Figure 2, Suppl. Table 3).

**Figure 2.**
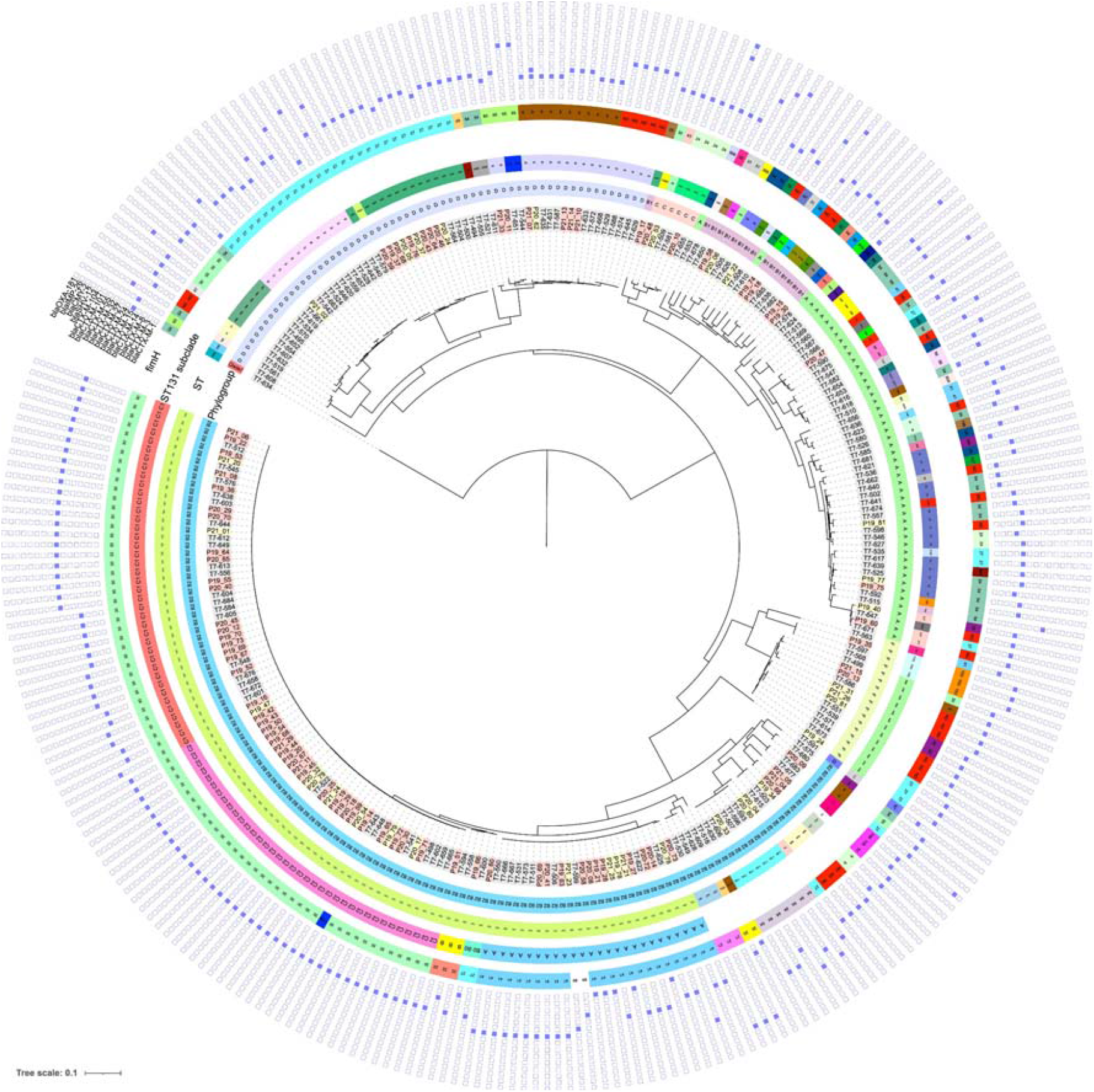
Maximum-likelihood phylogenetic tree based on core genome alignment of the genomes of ESBL-*E. coli* carriage isolates from Tromsø7 (labeled grey, n=166) and clinical isolates from NORM 2014 (blood isolates labeled with red and urine isolates with yellow, n=118). The innermost ring illustrates phylogroups, followed by a ring with sequence types (STs), a ring with ST131 subclades, and a ring with *fimH* types (ND, not detected). The heatmap shows presence (blue color) or absence (white) of ESBL gene variants.

At the phylogroup level, 52.4% of the ESBL-Ec carriage isolates belonged to either phylogroup A (26.5%) or D (25.9%), and 33.7% belonged to phylogroup B2 (Figure 2 and Figure 3, Suppl. Table 3). This contrasts with the clinical isolates where 64.4% (p<0.001) of the isolates belonged to phylogroup B2 and only 20.4% (p<0.001) belonged to phylogroup A (5.1%) and D (15.3%) combined.

**Figure 3.**
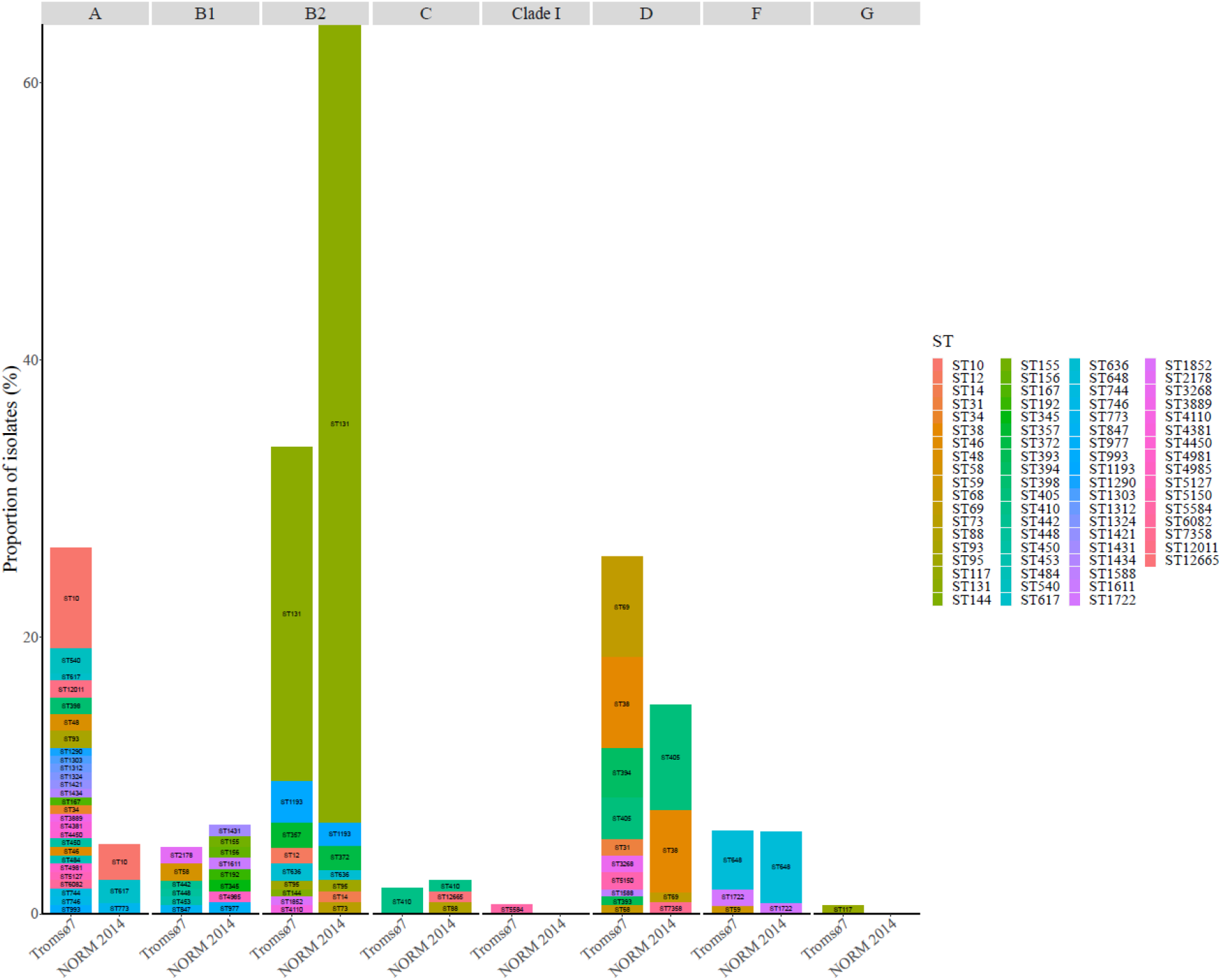
Phylogroup (labeled on the top) and sequence type (ST) distribution of ESBL-*E. coli* carriage isolates from Tromsø7 (n=166) and clinical isolates from NORM 2014 (n=118).

Carriage isolates had higher ST diversity (Figure 2 and 3, Suppl. Table 3) with Simpson’s diversity index 92.4% compared to clinical isolates (65.9%, p<0.001). We identified 58 different STs among the carriage isolates while the clinical isolates included 27 STs (Figure 2 and 3, Suppl. Table 3). ST131 (phylogroup B2) was the dominant ST in both collections, however, more prevalent among clinical isolates (57.6%, n=68) than carriage isolates (24.1%, n=40, p<0.001). Comparison of prevalence of ESBL-producing ST131 colonization vs. infection in our study results in a crude odds ratio (OR) for infection of 4.3 (2.57-7.13, p<0.001).

Within ST131, the multidrug resistant subclades C1 (alternatively referred to as *H*30-R) and C2 (*H*30-Rx) accounted for 67.5% in carriage and 73.5% clinical isolates (Figure 2, Suppl. Figure 1, Suppl. Table 3). The subclade C1 (47.5%, n=19) was the most prevalent among carriage isolates and C2 (42.6%, n=29) among clinical isolates. The difference in the proportion of C2 between clinical and carriage isolates was statistically significant (20.0%, n=8, p=0.017). For the overall ESBL-Ec population, crude OR for infection among C2 was 6.44 (2.82-14.68, p<0.001) (Suppl. Table 4).

Although non-significant, a higher proportion of subclades A, B and B0, less associated with antibiotic resistance, was observed among ST131 carriage isolates (32.5%) than clinical isolates (26.5%, p=0.508) (Suppl. Figure 1). However, with regard to the overall ESBL-Ec population, subclade A was associated with infection (carriage 6.0% vs. clinical 13.6%, OR 2.45, 1.07-5.60, p=0.034) (Suppl. Table 4).

The most prevalent STs in carriage isolates following ST131 were ST10 (7.2%, n=12, phylogroup A), ST69 (7.2%, n=12) and ST38 (6.6%, n=11, both phylogroup D) compared to ST405 (7.6%, n=9), ST38 (5.9%, n=7, both phylogroup D), and ST648 (5.1%, n=6, phylogroup F) in clinical isolates (Figure 2 and 3, Suppl. Table 3). We did not identify the common ExPEC lineage ST73^25,29^ among the carriage isolates but found one ST95^25,29^ (0.6%) and five isolates of the emerging ST1193^35,36^ (3.0%).

Using a single nucleotide polymorphism (SNP) cut-off of ≤17^37^, we detected two putative clusters (Suppl. Table 5) among five of 284 isolates. All five were carriage isolates. One ST357 cluster (6-9 SNP differences) consisted of three isolates and the other cluster of two ST131 isolates (4 SNP difference). We detected no clusters among the clinical isolates.

### Antimicrobial resistance and plasmid replicon content

CTX-M enzymes accounted for at least 97.0% of the ESBL phenotypes in both collections (Figure 2 and Figure 4, Suppl. Table 3). The most prevalent ESBL genes among both carriage and clinical isolates were *bla*CTX-M-15 (40.4%, n=67 vs. 61.0%, n=72, p=0.001), *bla*CTX-M-14 (20.5%, n=34 vs. 14.4%, n=17, p=0.188), and *bla*CTX-M-27 (19.9%, n=33 vs. 15.3%, n=18, p=0.321). One clinical isolate of ST38 harbored both *bla*CTX-M-15 and *bla*CTX-M-14. *Bla*CTX-M-3, *bla*CTX-M-8, *bla*CTX-M-32 and *bla*CTX-M-101 were exclusively detected in carriage isolates, whereas *bla*CTX-M-24 and *bla*CTX-M-104 were present only in clinical isolates. The remaining ESBL producers contained *bla*SHV-12 in 3.0% of the carriage and 1.7% of the clinical isolates. *Bla*TEM ESBL genes were not detected.

**Figure 4.**
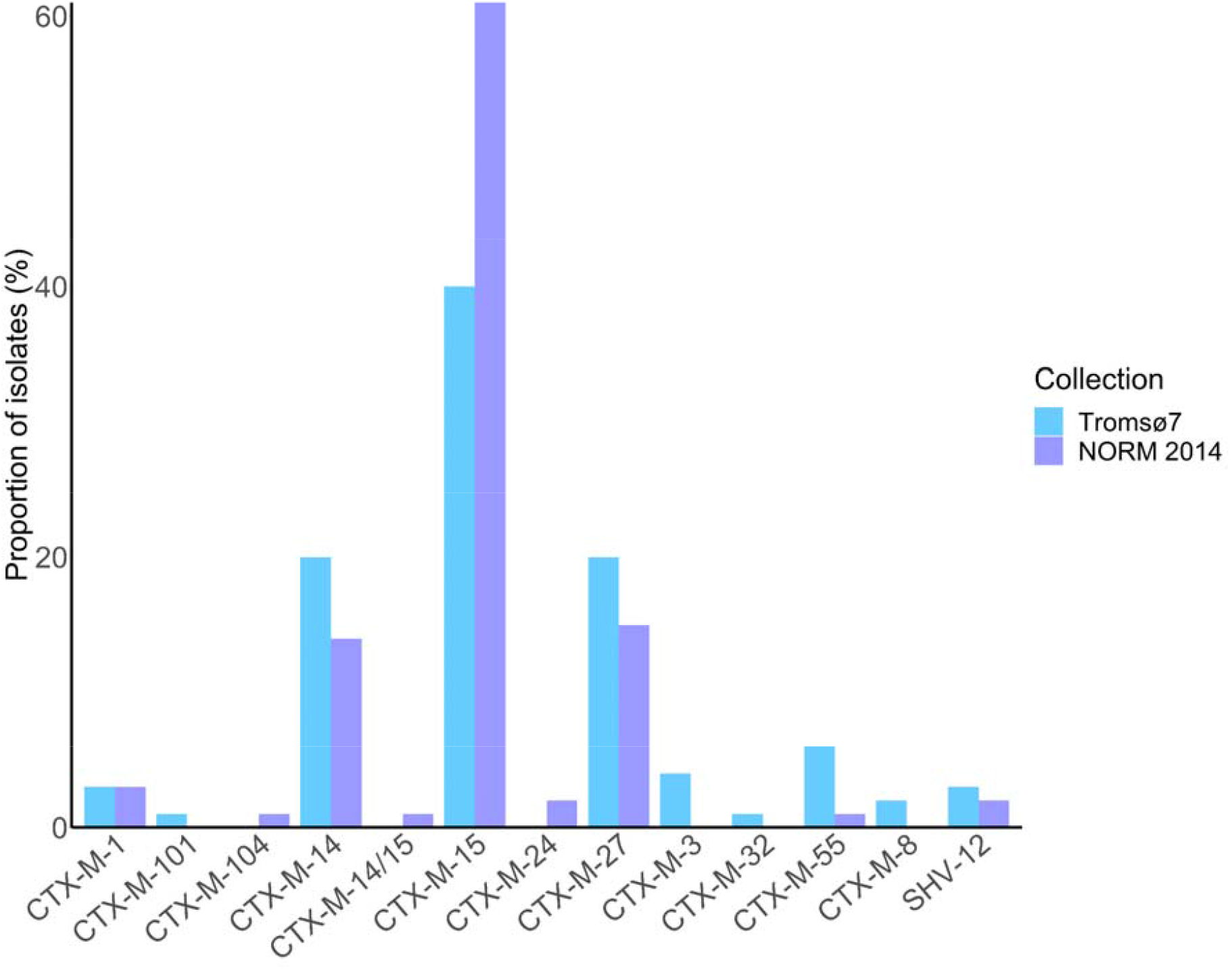
ESBL gene prevalence in ESBL-*E. coli* carriage isolates from the Tromsø7 study (n=166) and clinical isolates from NORM 2014 (n=118).

The clinical isolates showed an overall higher proportion of phenotypic resistance compared to the carriage isolates (Table 2, Suppl. Table 3). Only 2.4% of carriage isolates were phenotypically resistant to piperacillin-tazobactam compared to 31.8% (p<0.001) of clinical isolates. We also detected a high prevalence of phenotypic co-resistance to non-β-lactam antibiotics such as gentamicin, ciprofloxacin and trimethoprim-sulfamethoxazole both in clinical and carriage isolates. Isolates co-resistant to all these three antibiotic classes, accounted for 10.2% (17/166) of carriage and 33.1% (39/118, p<0.001) of clinical isolates.

**Table 2.**
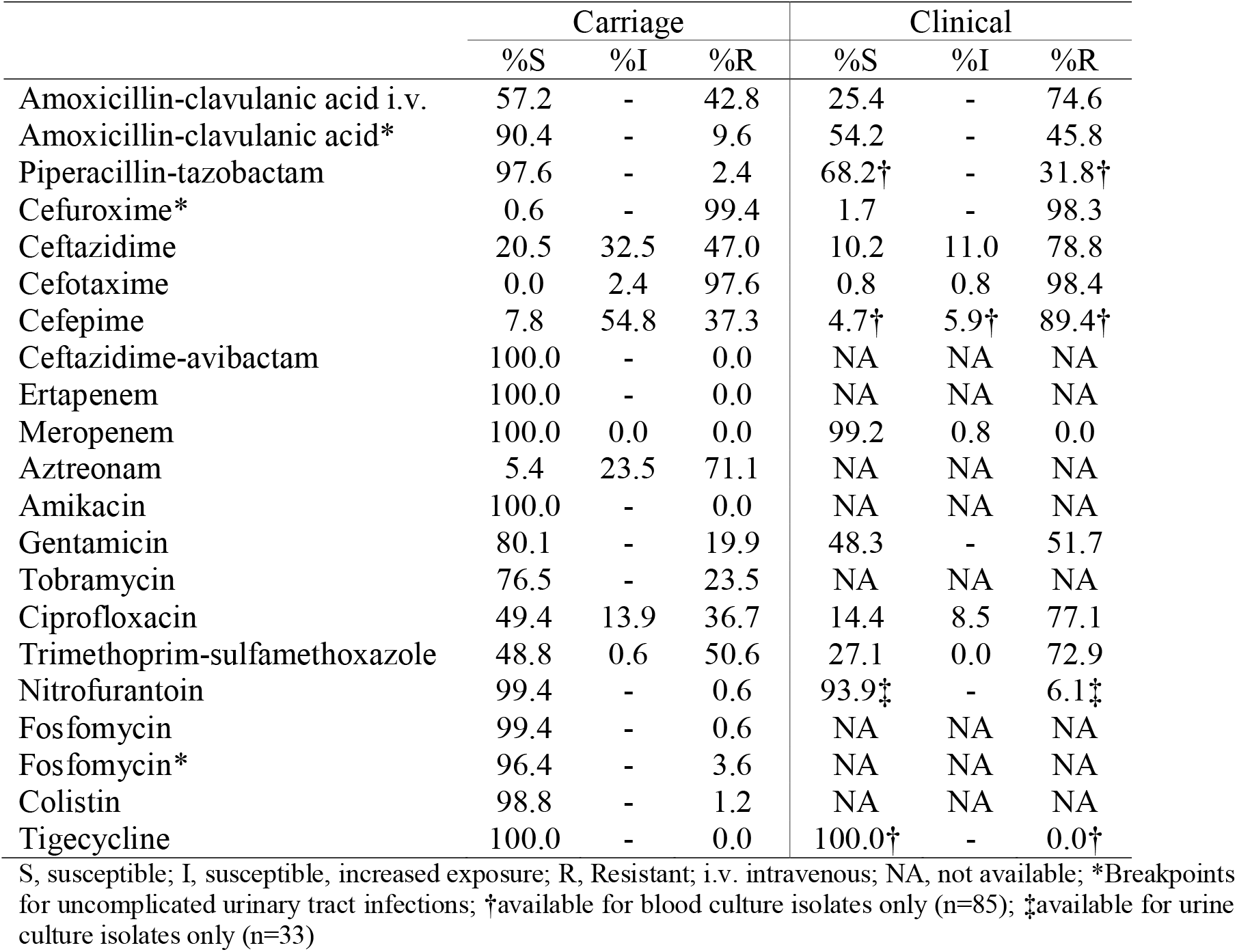
Susceptibility profile of ESBL-*E. coli* carriage isolates from Tromsø7 (n=166) and clinical isolates from NORM 2014 (n=118).

|  | Carriage |  |  | Clinical |  |  |
| --- | --- | --- | --- | --- | --- | --- |
|  | %S | %I | %R | %S | %I | %R |
| Amoxicillin-clavulanic acid i.v. | 57.2 | - | 42.8 | 25.4 | - | 74.6 |
| Amoxicillin-clavulanic acid* | 90.4 | - | 9.6 | 54.2 | - | 45.8 |
| Piperacillin-tazobactam | 97.6 | - | 2.4 | 68.2† | - | 31.8† |
| Cefuroxime* | 0.6 | - | 99.4 | 1.7 | - | 98.3 |
| Ceftazidime | 20.5 | 32.5 | 47.0 | 10.2 | 11.0 | 78.8 |
| Cefotaxime | 0.0 | 2.4 | 97.6 | 0.8 | 0.8 | 98.4 |
| Cefepime | 7.8 | 54.8 | 37.3 | 4.7† | 5.9† | 89.4† |
| Ceftazidime-avibactam | 100.0 | - | 0.0 | NA | NA | NA |
| Ertapenem | 100.0 | - | 0.0 | NA | NA | NA |
| Meropenem | 100.0 | 0.0 | 0.0 | 99.2 | 0.8 | 0.0 |
| Aztreonam | 5.4 | 23.5 | 71.1 | NA | NA | NA |
| Amikacin | 100.0 | - | 0.0 | NA | NA | NA |
| Gentamicin | 80.1 | - | 19.9 | 48.3 | - | 51.7 |
| Tobramycin | 76.5 | - | 23.5 | NA | NA | NA |
| Ciprofloxacin | 49.4 | 13.9 | 36.7 | 14.4 | 8.5 | 77.1 |
| Trimethoprim-sulfamethoxazole | 48.8 | 0.6 | 50.6 | 27.1 | 0.0 | 72.9 |
| Nitrofurantoin | 99.4 | - | 0.6 | 93.9‡ | - | 6.1‡ |
| Fosfomycin | 99.4 | - | 0.6 | NA | NA | NA |
| Fosfomycin* | 96.4 | - | 3.6 | NA | NA | NA |
| Colistin | 98.8 | - | 1.2 | NA | NA | NA |
| Tigecycline | 100.0 | - | 0.0 | 100.0† | - | 0.0† |

S, susceptible; I, susceptible, increased exposure; R, Resistant; i.v. intravenous; NA, not available; \*Breakpoints for uncomplicated urinary tract infections; †available for blood culture isolates only (n=85); ‡available for urine culture isolates only (n=33)

Phenotypic ciprofloxacin resistance was different for ST131 (70.0%) vs. non-ST131 (26.2%, p<0.001) among carriage isolates (Suppl. Table 6) in contrast to the clinical isolates (Suppl. Table 7). Two carriage isolates of ST484 and ST1324 (both phylogroup A) were phenotypically resistant to colistin and harbored *mcr*-1.1 and *mcr*-3.5, respectively. No isolates expressed clinical resistance against carbapenems. However, two clinical isolates of ST95 (phylogroup B2) and ST410 (phylogroup C) harbored *bla*IMP-26 and *bla*OXA-181, respectively. All isolates were susceptible to tigecycline.

Despite the overall higher proportion of phenotypic resistance among clinical isolates, the average number of different plasmid replicon-types per isolate (both 3.1/isolate) did not differ between the carriage and clinical collections. We identified 43 and 39 different plasmid replicon types among 97.1% (162/166) carriage and 95.0% (112/118) clinical isolates (Suppl. Table 3, Suppl. Figure 2). The most prevalent were IncFIB(AP001918) (carriage 22.7% vs. clinical 22.3%) followed by Col156 (carriage 11.1% vs. clinical 11.5%) and IncFIA (carriage 10.9% vs. clinical 17.7%, p=0.101). Replicon type Col156 and IncFIA were mainly detected in globally disseminated ExPEC clones (ST131, ST1193, ST648, ST69, ST405), whereas IncFIB(AP001918) was additionally frequently spread among other STs (Suppl. Table 3).

### ESBL-K. pneumoniae carriage isolates

We detected only four ESBL-Kp, each of a different ST (ST29, ST211, ST261 and ST2459) harboring *bla*CTX-M-15 (n=2), *bla*CTX-M-14 (n=1), or *bla*SHV-12 (n=1). None of the isolates were genomically assigned as hypervirulent.

## Discussion

Our study contributes to the knowledge of prevalence of, and factors associated with, ESBL-Ec/Kp gastrointestinal carriage in a general adult population, and the bacterial population structure of carriage isolates. The comparison to a contemporary collection of clinical ESBL-Ec isolates revealed differences in the population structure and the prevalence of phenotypic resistance between carriage and clinical isolates. Travel to Asia was identified as a major risk for ESBL-Ec gastrointestinal carriage.

An ESBL-Ec carriage prevalence of 3.3% (2.8-3.9) is lower but comparable to previous community-based data from Europe including Sweden 4.4% (3.5-5.3, n=2,134, data collected 2012-2013)^13^, the Netherlands 4.5% (3.9-5.1, n=4,177, 2014-2016)^14^ and a Norwegian study 4.9% (2.7-8.1, n=284, 2014-2016)^17^ using similar screening approaches.

We identified significant differences in the ESBL-Ec population structure between the community and clinical isolates. The globally disseminated phylogroup B2 clone ST131 has been identified as a key contributor to the increase in ESBL prevalence^38^ and in a longitudinal study of *E. coli* bloodstream isolates we identified ST131 to be the single largest contributor to the increase in prevalence of ESBL-Ec in Norway.^25^ We also observed the predominance of ST131 in both our collections. However, the proportion is significantly lower in carriage isolates due to lower numbers of the multidrug resistant subclade C2. Moreover, the carriage population had a higher proportion of phylogroup A, associated with asymptomatic intestinal carriage in humans^39,40^ and a significantly greater ST diversity overall, compared to the clinical isolates. These observations indicate that acquisition of ESBL genes frequently occur in a variety of *E. coli* lineages colonizing the gut. However, there are differences in the colonization potential of *E. coli* lineages and the risk of invasive infection by ESBL-Ec which seems to be clone dependent.^38,41^ The higher odds for infection that we detected for ST131 is similar to that of the Swedish study (AOR 3.4, 1.8-6.4) indicating a higher pathogenicity potential of ST131 compared to commensal *E. coli* lineages of phylogroup A, such as ST10.^13^ Moreover, we found that ST131 subclade A, previously reported with less resistance, and the multidrug resistant subclade C2 had higher odds for infection, and this may contribute to the sustained establishment of these subclades among bloodstream infections in Norway.^25^

Assuming a 100% colonization rate of *E. coli*, the large proportion of STs notorious as common causes of extra-intestinal clinical infections (e.g. ST131, ST405, ST38 and ST648)^25,27,28^ could at least partly explain the higher prevalence of ESBL among *E. coli* causing bloodstream infections in Norway (5.8% in 2016)^42^ compared to the carriage prevalence of 3.3% identified here. We also observed emerging clones such as ST1193 which appears to have disseminated rapidly worldwide over the last decade.^35,36^ The low prevalence of ESBL-Kp gastrointestinal carriage (0.08%) is consistent with previous community based reports.^9,14,17^

The identification of clusters with closely related isolates could indicate putative clonal spread. This could include within-household and social network transmission or nosocomial spread. However, we did not have access to epidemiological data to examine this further.

In line with other studies, we found a strong association between ESBL-Ec carriage and travel to Asian regions.^13-15,17,22^ This supports the current patient screening recommendations for ESBL-producing gram-negative bacteria after hospital stay abroad in the past year before hospital admission in Norway.^43^ In contrast to a Swedish and a Dutch study,^15,44^ we did not identify travelers’ diarrhea as a risk factor for ESBL-Ec carriage. However, our study was not designed to specifically investigate international travelers but rather focused on risk factors in the general adult population.

We found no association between ESBL-Ec gut carriage and factors such as hospitalization, antibiotic use, and acid suppressive medication, and conflicting results have been detected in previous studies.^22,45^ Hospitalization as a risk factor has mainly been reported from studies investigating patients with ESBL-E infections.^23,46^ In line with most studies that assessed risk factors regarding ESBL-E carriage in individuals in the community, we did not identify hospitalization as an independent risk factor.^13,22,44^ This may be due to the increased ESBL prevalence not only in hospitals, but also in the community over the last decades and implies that boundaries have become blurred between those two settings.^9,22,47,48^

The nonsignificant effect of antibiotic use is mainly due to limitations of the drug variable in our study which is based on self-reported data, including topical antibiotics and covers only the last two weeks before self-sampling. As antibiotic use has been found as a risk factor for resistance in many other studies,^14,15,22^ we cannot rule out that antibiotic use plays a role in ESBL-E carriage. There are reports identifying an association between the use of gastric acid suppressive medication and intestinal colonization or infections with ESBL-E.^18,21,23,33,34,46^

However, a Dutch study comparable to ours did not find an association between proton pump inhibitor use and ESBL-E carriage in the overall analysis.^14^

Interestingly, we found a possible association between Kp and ESBL-Ec carriage. An association between ESBL-E carriage and vancomycin-resistant enterococci (VRE) has been described previously^7,49^, and also that VRE colonization is significantly associated with Kp colonization among intensive care unit patients.^50^ These associations warrant further investigations to assess if a common set of risk factors for carriage of different clinically important pathogens can be identified.

An important strength of our study is the non-selective recruitment from the official population-registry, and the high participation (87%) compared to 18.3% and 18.8% in comprehensive studies from the Netherlands^14^ and Sweden^13^, respectively. It is a limitation that we only captured the general population 40 years and older. However, other studies have not found an association between age and ESBL-E carriage.^13,14^ Moreover, more extensive data on drug use would have strengthened the analyses. Additionally, the genomic diversity of carriage isolates is likely to be underestimated due to the isolation and sequencing of only one colony per fecal sample. Although, we compared local carriage isolates with a national collection of clinical ESBL-Ec, there was no discernible spatiotemporal spread or phylogenetic structure within Norway according to a nationwide genomic study on *E. coli* causing bloodstream infections.^25^

## Conclusions

The prevalence of ESBL-Ec carriage in a general adult urban Norwegian population was low reflecting the relatively low prevalence of ESBL-Ec in clinical isolates. Travel to Asia was the only independent risk factor for ESBL-Ec carriage and should be considered in terms of screening recommendations before hospital admission. The differences in ESBL-Ec populations between carriage and clinical isolates indicating a higher risk of infection dependent on the ESBL-Ec clone warrants further investigations to elucidate if risk stratification should include determination of the bacterial genetic background.

## Subjects and Methods

### Study population and design

Our study sample was drawn from Tromsø7, the last of seven cross-sectional health surveys conducted between 1974 and 2016 in Tromsø municipality, Norway (https://uit.no/research/tromsostudy). Tromsø is representative of a Northern European, urban population.^31^ Tromsø7 (March 2015-October 2016) included questionnaires and two clinical visits (https://uit.no/research/tromsostudy/project?pid=708909). Unique national identity numbers from the official population-registry were used to invite all citizens >40 years (n=32,591). Sixty-five percent (n=21,083, 11,074 women) attended the first clinical visit in the study (Figure 5).

**Figure 5.**
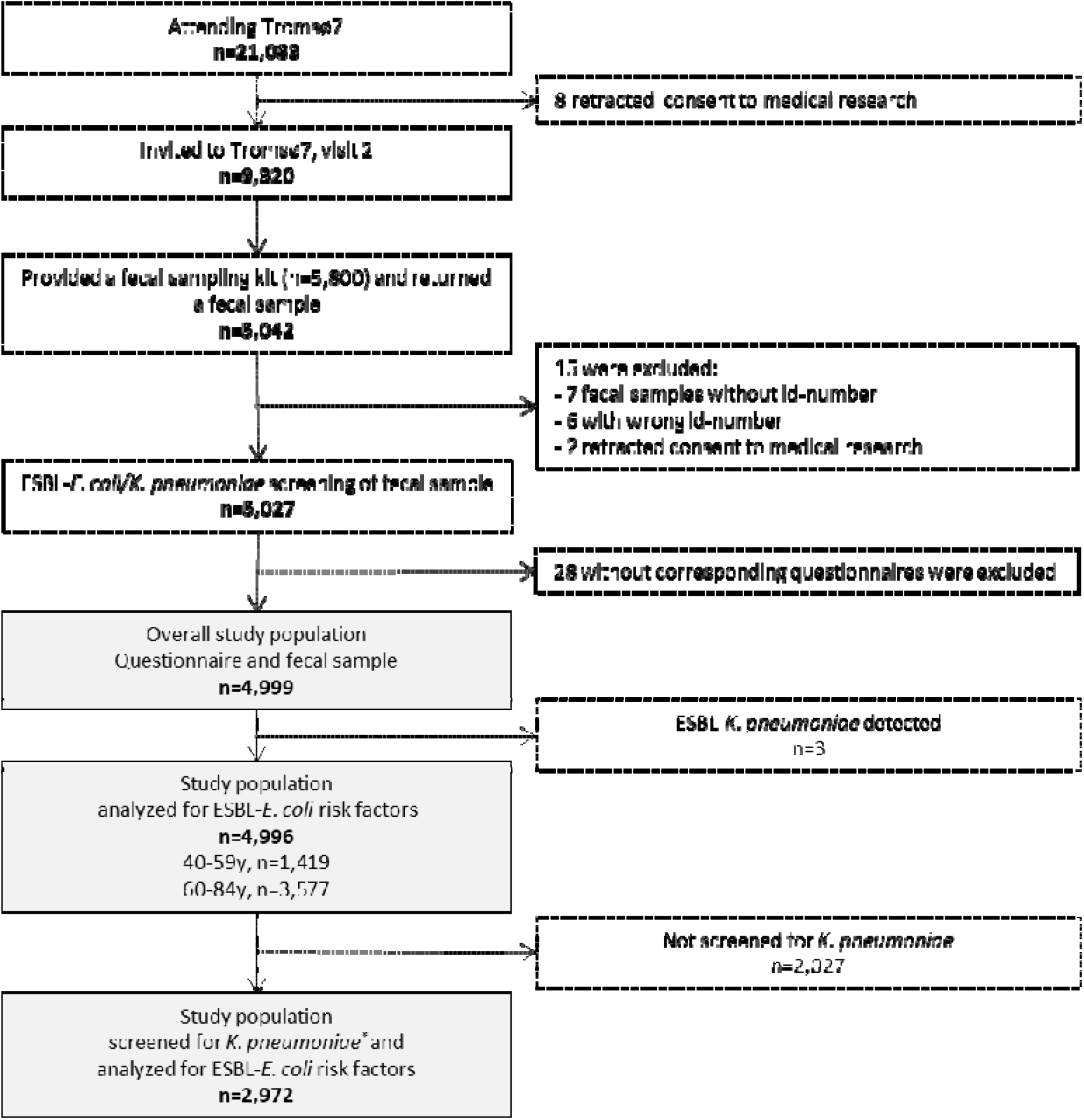
Flow diagram of the study population, Tromsø7, 2015-2016. *Study population screened for *K. pneumoniae* carriage is described in.^32^

A selection of 9,320 persons attending the first visit, were invited for a second visit, also including 3,154 former Tromsø Study participants not already included in the random selection process, which was required for other clinical research purposes. From March 2015 to March 2016, 5,800 participants at the first visit were consecutively provided a fecal self-sampling kit. Participants collected fecal material using nylon-flocked ESwab 490CE.A (Copan, Brescia, Italy). In total, 87% (n=5,042) returned a sample either at the second visit, or by mail to the laboratory.

All 5,042 fecal samples were screened for the presence of ESBL-producing Ec and Kp via selective culture (see below). All participants completed two self-administered structured questionnaires on socio-demographics, smoking, alcohol use, hospitalization, drug use, and travel abroad. We excluded 13 participants with wrong or missing sample identification number, two retracting consent to medical research, and 28 with incomplete questionnaires for a final study population of 4,999 participants (Figure 5). We analyzed the association between ESBL-Ec gastrointestinal carriage and different risk factors in 4,996 participants (Figure 5 and Table 1). Next, we investigated the association between ESBL-Ec carriage and Kp gastrointestinal carriage among 2,972 participants additionally screened for Kp in our previous study,^32^ irrespective of resistance (Figure 5, Supplementary Table 2).

### Isolation of ESBL-producing E. coli and K. pneumoniae

We added 200 µl 85% glycerol to the ESwab tubes upon arrival at the local microbiological laboratory, and stored the samples at −80°C. From the thawed media, 100 µl were plated onto CHROMagar™ ESBL (CHROMagar, Paris, France) and incubated for 48 hours at 37°C. Pink, purple and blue colonies suspected of being ESBL-producing Ec or *Klebsiella* spp. were identified using mass spectrometry (MALDI-TOF, Bruker Daltonics, Bremen, Germany). The first colony identified as either Ec, *K. pneumoniae* or *K. variicola* from each sample was kept and further analyzed. All samples were plated on cysteine lactose electrolyte deficient agar (MAST Group, Bootle, UK) to assess growth of fecal flora and validity of the samples.

### K. pneumoniae isolation

The screening strategy and isolation procedure for Kp, is described in detail elsewhere^32^. Briefly, we plated and screened the fecal samples onto the selective SCAI (Simons citrate agar with inositol; both Sigma-Aldrich, Darmstadt, Germany) medium and identified suspected colonies using MALDI-TOF.

### Antimicrobial susceptibility testing and phenotypic ESBL-identification

Susceptibility testing was performed according to the EUCAST broth microdilution method for carriage isolates, disc diffusion method^51^ for the clinical isolates and both interpreted using the EUCAST 2023 breakpoint table (https://eucast.org/). For the confirmation of ESBL-producing Ec and Kp, we followed the EUCAST algorithm for phenotypic detection of ESBLs using the BD BBL™ combination disk test (Becton Dickinson and Company, Sparks, USA).

#### Genomic sequencing and bioinformatic analysis

Genomic DNA from 166 Ec of the Tromsø7 collection was extracted with the MagNA Pure 96 system (Roche Applied Science, Mannheim, Germany) and sequencing libraries were prepared according to the Nextera Flex sample preparation protocol (Illumina, San Diego, CA, USA). Samples were sequenced on the Illumina MiSeq platform to generate 300 bp paired-end reads. All reads were trimmed with TrimGalore v0.6.4 and assembled with Unicycler v0.4.8 including SPAdes v3.13.0.^52–54^ STs were assigned using the multi locus sequence type (MLST) software *E*.*coli* scheme v2.19.0 and Enterobase.^55–57^ AMRFinderPlus v3.10.16 was used to determine resistance genes among the Ec isolates.^58^ Plasmid replicons were identified using Abricate v1.0.1 including the PlasmidFinder 2021-Mar-27 database.^59,60^ Phylogroup assignment was based on ClermonTyping v20.03 (Mar. 2020).^61^ The *fimH* type was identified using FimTyper v1.0 and BLAST+ v2.12.0.^62,63^ Regarding the *Klebsiella* isolates, Kleborate v2.0.0 was used to determine species identification, ST and acquired genes encoding virulence or antibiotic resistance.^64,65^

The clinical ESBL-producing Ec included 118 isolates out of 123 representing all ESBL-Ec isolates collected in 2014^66^, as part of the yearly surveillance program in NORM. Genome sequencing of five isolates were unsuccessful. Before sequencing, the NORM 2014 isolates were stored at −80°C and then sent to GATC Biotech AG (part of Eurofins Genomics/Eurofins Scientific) in Germany for DNA isolation and WGS. Raw reads were trimmed using Trimmomatic v0.39 and assembled with SPAdes v3.15.0.^67,68^ Contigs shorter than 200 bp were discarded. The genomic data were analyzed as described above.

### Phylogenetic and population structure analysis of ESBL-E. coli

We used Prokka v.1.14.6^69^ to annotate genomes and snippy v.4.6.0^70^ to map the sequence reads to the ST131 Ec EC958 chromosome [HG941718.1] (https://www.ncbi.nlm.nih.gov/nuccore/HG941718.1) to create the core genome alignment. We used snp-dists v.0.8.2, (https://github.com/tseemann/snp-dists/) to create the SNP distance matrix from the core genome alignment. We used a 17 SNP cut-off to define similarity between two genomes and to identify probable ESBL-Ec transmission events among cases.^37^ To assess the phylogenetic relatedness, we used the core SNP alignment to infer a maximum-likelihood tree using RAxML v.8.2.8 with the GTR + Gamma rate model and 100 rapid bootstraps visualized in iTol (v6.5.2).^71,72^ ST131 subclades were determined based on subclade specific SNPs and *fimH* alleles, subclade membership was corrected when assignment based on SNP profile of sporadic isolates did not fit with the phylogenetic distribution of clades.^25,30,62,73^

### Statistical analysis

Our primary analyses were multivariable logistic regression models, performed in two overlapping study populations with outcome variable ESBL-Ec gastrointestinal carriage using SPSS v.26.0 (SPSS, Inc., Chicago, IL, USA). First, we analyzed factors associated with ESBL-Ec gastrointestinal carriage among 4,996 participants (Table 1). Next, in 2,972 participants also screened for Kp,^32^ we performed a multivariable logistic regression analysis including Kp carriage as an additional explanatory variable (Suppl. Table 2). Explanatory variables were selected with the help of a directed acyclic graph constructed using DAGitty v3.0 (Suppl. Figure 3 and 4).^74^ All explanatory variables were kept in the fully adjusted models. Multicollinearity between the entered variables was assessed calculating the variance inflation factor (VIF) and tolerance statistic. Multicollinearity was not a problem with VIF>10 and tolerance statistic <0.2.^75^ The strength of the associations was examined by calculating AORs with 95% CI. Two-sided p-values <0.05 were considered statistically significant. The prevalence of ST131 among carrier and clinical isolates was compared calculating the OR with 95% CI using logistic regression in SPSS. The comparison of proportions was assessed using χ^2^-test.

## Supporting information

Supplementary Table 3. Genome characteristics of ESBL-E. coli from carriage isolates and clinical isolates

## Data Availability

Bacterial genome data (raw Illumina reads) are publicly available in NCBI under BioProject PRJEB53319 (NORM 2014 collection) and PRJEB57251 (Tromsø7 collection). This study is based on data owned by a third party (The Tromsø Study, Department of Community Medicine, UiT The Arctic University of Norway). Confidentiality requirements according to Norwegian law prevents sharing of individual patient level data in public repositories. Application of legal basis and exemption from professional secrecy requirements for the use of personal health data in research may be sent to a regional committee for medical and health research ethics (https://rekportalen.no/). The authors gained access to the data through the Tromsø Study application process. Guidelines on how to access the data are available at the website https://uit.no/research/tromsostudy. All enquiries about the Tromsø Study should be sent by e-mail to. All the questionnaire variables are published in the NESSTAR program system and results can be viewed online: http://tromsoundersokelsen.uit.no/tromso/.

## Ethics

The study was approved by the Regional Committee for Medical and Health Research Ethics, North Norway (REC North reference: 2016/1788 and 2014/940) and the Data Protection Officer at University Hospital of North Norway (reference: 2019/4264). The study complied with the Declaration of Helsinki. All participants in Tromsø7 signed an informed consent form prior to participation.

## Acknowledgements

We are grateful for technical assistance from Bjørg Haldorsen, Bettina Aasnæs and Ellen Josefsen in organizing the collection of fecal samples in the laboratory, Eva Bernhoff and Ragna-Johanne Bakksjø for performing WGS, Marit Andrea Klokkhammer Hetland for initial sequence analysis of carriage isolates. Rod Wolstenholme for figure editing. We thank Sigma2, the national provider of e-infrastructure, for access to High-Performance Computing and large-scale data storage (Project ID: NN9794K).

Collaborators forming the Norwegian *E. coli* ESBL Study Group include: Nina Handal (Akershus University Hospital), Elisabeth Sirnes (Førde Hospital), Liv Jorunn Hafne (Haugesund Hospital), Paul Christoffer Lindemann (Haukeland University Hospital), Lovise Marie Norgaard (Innlandet Hospital), Kyriakos Zaragkoulias (Levanger Hospital), Einar Nilsen (Molde and Ålesund Hospital), Hege Elisabeth Larsen (Nordland Hospital), Marcela Pino (Oslo University Hospital, Rikshospitalet), Nils Olav Hermansen (Oslo University Hospital, Ullevål), Iren H. Löhr (Stavanger University Hospital), Aleksandra Jakovljev (St. Olav University Hospital), Ståle Tofteland (Sørlandet Hospital), Kristina Papp (Unilabs Telelab), Gunnar Skov Simonsen (University Hospital of North Norway), Åshild Marvik (Vestfold Hospital), Nadine Durema Pullar (Vestre Viken, Bærum Hospital), Roar Magne Bævre-Jensen (Vestre Viken, Drammen Hospital), Anita Kanestrøm (Østfold Hospital).

## Author contributions

ØS conceptualized and acquired funding for the study in collaboration with AS, KG and IHL. ØS was responsible for organizing the collection of fecal samples. LLEA did the screening of fecal samples and phenotypic testing. NR, KS, LS and KG conceptualized and conducted the epidemiological analysis of risk factors. IHL organized WGS of carriage isolates. GSS and the Norwegian *E. coli* ESBL Study Group provided data and clinical isolates. DJB, AKP, NR and ØS analyzed the WGS data. NR, ØS, KS and KG prepared first manuscript draft. All authors contributed to review and editing of the manuscript and approved the final version.

## Data Availability

Bacterial genome data (raw Illumina reads) are publicly available in NCBI under BioProject PRJEB53319 (NORM 2014 collection) and PRJEB57251 (Tromsø7 collection). This study is based on data owned by a third party (The Tromsø Study, Department of Community Medicine, UiT The Arctic University of Norway). Confidentiality requirements according to Norwegian law prevents sharing of individual patient level data in public repositories. Application of legal basis and exemption from professional secrecy requirements for the use of personal health data in research may be sent to a regional committee for medical and health research ethics (https://rekportalen.no/). The authors gained access to the data through the Tromsø Study’s application process. Guidelines on how to access the data are available at the website https://uit.no/research/tromsostudy. All enquiries about the Tromsø Study should be sent by e-mail to. All the questionnaire variables are published in the NESSTAR program system and results can be viewed online: http://tromsoundersokelsen.uit.no/tromso/.

