## Supplementary material, Tables and Figures for "Community carriage of ESBL-producing *Escherichia coli* and *Klebsiella pneumoniae*: A cross-sectional study of risk factors and comparative genomics of carriage and clinical isolates"

*^g^Collaborators forming the Norwegian E. coli ESBL Study Group are listed in the Acknowledgement.*

*^h^Department of Microbiology and Infection Control, Akershus University Hospital, Nordbyhagen, Norway*

*^i^Division of Medicine and Laboratory Sciences, Institute of Clinical Medicine, University of Oslo, Oslo, Norway*

^#^ equal contribution

*Corresponding author: Niclas Raffelsberger, Correspondence address: Department of Microbiology and Infection Control, University Hospital of North Norway, N-9038 Tromsø, Norway.

**Table of contents**

Supplementary Table 1. Characteristics of the overall study population (n=4,999) and the ESBL-*E. coli* study population (n=4,996) in Tromsø7, 2015-2016.

Supplementary Table 2. Association between ESBL-producing *E. coli* and *K. pneumoniae* species complex gastrointestinal carriage among 2,972 participants in Tromsø7.

Supplementary Table 3. Genome characteristics of ESBL-*E. coli* from carriage isolates from Tromsø7 (n=166) and clinical isolates from NORM 2014 (n=118) isolates, (provided as Excel-file).

Supplementary Figure 1. ST131 clade distribution among ESBL-*E. coli* carriage isolates from Tromsø7 (n=40) and clinical isolates from NORM 2014 (n=68).

Supplementary Table 4. Comparison of ESBL-*E. coli* ST131 subclade prevalence between carriage isolates from Tromsø7 (n=166) and clinical isolates from NORM 2014 (n=118).

Supplementary Table 5. SNP distances among two putative ESBL-producing *E. coli* clusters identified using a ≤17 SNP cut-off.

Supplementary Table 6. Susceptibility profile of carriage isolates of ESBL-*E. coli* ST131 (n=40) and non-ST131 (n=126) from Tromsø7.

Supplementary Table 7. Susceptibility profile of clinical isolates of ESBL-*E. coli* ST131 (n=68) and non-ST131 (n=50) from NORM 2014

Supplementary Figure 2. Replicon type distribution among ESBL-*E. coli* from carriage isolates from Tromsø7 (n=166) and clinical isolates from NORM 2014 (n=118).

Supplementary Figure 3. Directed acyclic graph (DAG) for the first multivariable logistic regression model.

Supplementary Figure 4. Directed acyclic graph (DAG) for the second multivariable logistic regression model.

**Supplementary Table 1.** Characteristics of the overall study population (n = 4,999) and the ESBL-*E. coli* study population (n = 4,996) in Tromsø7, 2015-2016.

|  | Overall study population | | ESBL-*E. coli* study population | |
| --- | --- | --- | --- | --- |
| Characteristics | N | % | N | % |
| Sex |  |  |  |  |
| Men | 2,296 | 45.9 | 2,293 | 45.9 |
| Women | 2,703 | 54.1 | 2,703 | 54.1 |
| Age (years) |  |  |  |  |
| 40-49 | 605 | 12.1 | 605 | 12.1 |
| 50-59 | 814 | 16.3 | 814 | 16.3 |
| 60-69 | 2,128 | 42.6 | 2,127 | 42.6 |
| 70-84 | 1,452 | 29.0 | 1,450 | 29.0 |
| Household income |  |  |  |  |
| Low | 2,015 | 42.7 | 2,013 | 42.7 |
| High^a^ | 2,699 | 57.3 | 2,698 | 57.3 |
| Current daily smoking |  |  |  |  |
| No | 4,360 | 88.0 | 4,357 | 88.0 |
| Yes | 595 | 12.0 | 595 | 12.0 |
| Alcohol consumption frequency |  |  |  |  |
| Never to ≤monthly | 1,624 | 32.7 | 1,624 | 32.7 |
| 2-4/month or 2-3/week | 1,820 | 36.6 | 1,818 | 36.6 |
| ≥4/week | 1,527 | 30.7 | 1,526 | 30.7 |
| Hospitalization past 12 months |  |  |  |  |
| No | 4,344 | 88.0 | 4,341 | 88.0 |
| Yes | 593 | 12.0 | 593 | 12.0 |
| Antibiotic use past 14 days^b^ |  |  |  |  |
| No | 4,832 | 96.9 | 4,831 | 96.9 |
| Yes | 155 | 3.1 | 153 | 3.1 |
| Acid suppressive medication last 4 weeks |  |  |  |  |
| No | 3,762 | 79.7 | 3,760 | 79.7 |
| ≤weekly | 403 | 8.5 | 402 | 8.5 |
| Every week, but not daily | 260 | 5.5 | 260 | 5.5 |
| Daily | 297 | 6.3 | 297 | 6.3 |
| Travel abroad past 12 months^c^ |  |  |  |  |
| No | 2,145 | 44.5 | 2,145 | 44.5 |
| Other regions (excl. Asia) | 2,214 | 45.9 | 2,212 | 45.9 |
| Asia exclusively or Asia + other regions | 462 | 9.6 | 461 | 9.2 |
| Traveler`s diarrhea past 12 months^d^ |  |  |  |  |
| No | 4,724 | 96.6 | 4,721 | 96.6 |
| Yes | 166 | 3.4 | 166 | 3.4 |

^a^ ≥551 000 NOK (€ 53 976/year as per June 2022)

^b^ Have you taken any antibiotics (tablets or oral suspensions, nasal ointments, eye drops or eye ointment) during the past 14 days?

^c^ Traveled outside the Nordic countries >1 week duration in the past 12 months.

^d^ For each travel abroad past 12 months, the participants were asked if they did experience diarrhea in connection with the travel.

**Supplementary Table 2.** Association between ESBL-producing *E. coli* and *K. pneumoniae* species complex (SC) gastrointestinal carriage among 2,972 participants in Tromsø7.

| Characteristics | % (ESBL- *E. coli*) | 95% CI | n (ESBL- *E. coli*) | N | AOR | 95% CI | p-value |
| --- | --- | --- | --- | --- | --- | --- | --- |
| *K. pneumoniae* SC gastrointestinal carriage |  |  |  |  |  |  | 0.055 |
| No | 2.7 | 2.1-3.4 | 67 | 2,488 | 1.00 |  |  |
| Yes | 4.8 | 3.2-7.0 | 23 | 484 | 1.66 | 0.99-2.79 |  |

ESBL, extended spectrum β-lactamase; N, denominator; AOR, adjusted odds ratio; CI, confidence interval

AOR adjusted for age, hospitalization past 12 months, antibiotic use past 14 days, acid suppressive medication past 4 weeks, travel abroad past 12 months, traveler`s diarrhea past 12 months and *K. pneumoniae* SC gastrointestinal carriage.

The multivariable model includes 2,650 participants with complete information on all variables.

**Supplementary Table 3.** Genome characteristics of ESBL-*E. coli* carriage isolates from Tromsø7 (n=166) and clinical isolates from NORM 2014 (n=118) isolates, (provided as Excel-file).

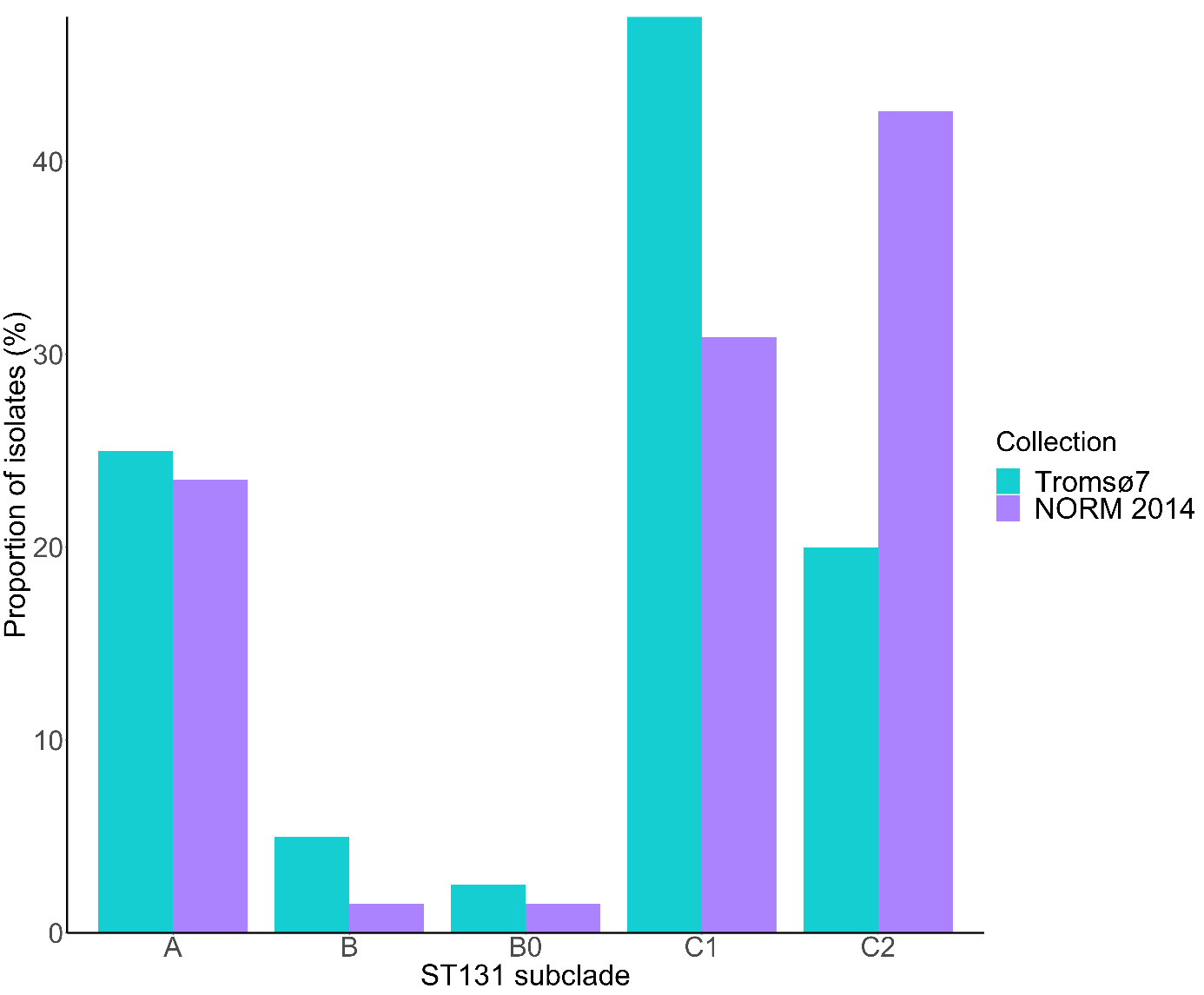
**Supplementary Figure 1.** Subclade distribution among ST131 ESBL-*E. coli* carriage isolates from Tromsø7 (n=40) and clinical isolates from NORM 2014 (n=68).

**Supplementary Table 4.** Comparison of ESBL-*E. coli* ST131 subclade prevalence between carriage isolates from Tromsø7 (n=166) and clinical isolates from NORM 2014 (n=118).

| ST131 subclade | Tromsø7 % (n) | NORM 2014 % (n) | OR | 95%CI | p-value |
| --- | --- | --- | --- | --- | --- |
| A | 6.0 (10) | 13.6 (16) | 2.45 | 1.07-5.60 | 0.034 |
| B | 1.2 (2) | 0.8 (1) | 0.70 | 0.06-7.82 | 0.773 |
| B0 | 0.6 (1) | 0.8 (1) | 1.41 | 0.09-22.78 | 0.809 |
| C1 | 11.4 (19) | 17.8 (21) | 1.67 | 0.86-3.28 | 0.132 |
| C2 | 4.8 (8) | 24.6 (29) | 6.44 | 2.82-14.68 | <0.001 |

OR, odds ratio; CI, confidence interval

**Supplementary Table 5.** SNP distances among two putative ESBL-producing *E. coli* clusters identified using a ≤17 SNP cut-off.

| Cluster | SNP distance | Sample ID | Collection | Sample material | ST | ST131 subclade |
| --- | --- | --- | --- | --- | --- | --- |
| 1 | 4 | T7-498 | Tromsø7 | Feces | ST131 | C1 |
|  |  | T7-602 | Tromsø7 | Feces |  |  |
| 2 | 6-9 | T7-530  T7-549 | Tromsø7 | Feces | ST357 | - |
|  |  | T7-628 | Tromsø7 | Feces |  |  |

SNP, single nucleotide polymorphism

**Supplementary Table 6.** Susceptibility profile of carriage isolates of ESBL-*E. coli* ST131 (n=40) and non-ST131 (n=126) from Tromsø7.

|  | ST131 | | | non-ST131 | | |
| --- | --- | --- | --- | --- | --- | --- |
|  | %S | %I | %R | %S | %I | %R |
| Amoxicillin-clavulanic acid i.v. | 57.5 | - | 42.5 | 57.1 | - | 42.9 |
| Amoxicillin-clavulanic acid* | 90.0 | - | 10.0 | 90.5 | - | 9.5 |
| Piperacillin-tazobactam | 97.5 | - | 2.5 | 97.6 | - | 2.4 |
| Cefoxitin | 100.0 | - | 0.0 | 88.1 | - | 11.9 |
| Cefuroxime* | 0.0 | - | 100.0 | 0.8 | - | 99.2 |
| Ceftazidime | 17.5 | 42.5 | 40.0 | 21.4 | 29.4 | 49.2 |
| Cefotaxime | 0.0 | 2.5 | 97.5 | 0.0 | 2.4 | 97.6 |
| Cefepime | 10.0 | 60.0 | 30.0 | 7.1 | 53.2 | 39.7 |
| Ceftazidime-avibactam | 100.0 | - | 0.0 | 100.0 | - | 0.0 |
| Ertapenem | 100.0 | - | 0.0 | 100.0 | - | 0.0 |
| Meropenem | 100.0 | 0.0 | 0.0 | 100.0 | 0.0 | 0.0 |
| Aztreonam | 5.0 | 30.0 | 65.0 | 5.6 | 21.4 | 73.0 |
| Amikacin | 100.0 | - | 0.0 | 100.0 | - | 0.0 |
| Gentamicin | 70.0 | - | 30.0 | 83.3 | - | 16.7 |
| Tobramycin | 65.0 | - | 35.0 | 80.2 | - | 19.8 |
| Ciprofloxacin | 20.0 | 10.0 | 70.0 | 58.7 | 15.1 | 26.2 |
| Trimethoprim-sulfamethoxazole | 42.5 | 0.0 | 57.5 | 50.8 | 0.8 | 48.4 |
| Nitrofurantoin | 97.5 | - | 2.5 | 100.0 | - | 0.0 |
| Fosfomycin | 100.0 | - | 0.0 | 99.2 | - | 0.8 |
| Fosfomycin* | 97.5 | - | 2.5 | 96.0 | - | 4.0 |
| Colistin | 100.0 | - | 0.0 | 98.4 | - | 1.6 |
| Tigecycline | 100.0 | - | 0.0 | 100.0 | - | 0.0 |

S, susceptible; I, susceptible, increased exposure; R, resistant; i.v. intravenous; *Breakpoints for uncomplicated urinary tract infections;

**Supplementary Table 7.** Susceptibility profile of clinical isolates of ESBL-*E. coli* ST131 (n = 68) and non-ST131 (n = 50) from NORM 2014

|  | ST131 | | | non-ST131 | | |
| --- | --- | --- | --- | --- | --- | --- |
|  | %S | %I | %R | %S | %I | %R |
| Amoxicillin-clavulanic acid i.v. | 30.9 | - | 69.1 | 18.0 | - | 82.0 |
| Amoxicillin-clavulanic acid* | 54.4 | - | 45.6 | 54.0 | - | 46.0 |
| Piperacillin-tazobactam† | 70.4 | - | 29.6 | 64.5 | - | 35.5 |
| Cefuroxime* | 0.0 | - | 100.0 | 4.0 | - | 96.0 |
| Ceftazidime | 13.2 | 11.8 | 75.0 | 6.0 | 10.0 | 84.0 |
| Cefotaxime | 0.0 | 0.0 | 100.0 | 2.0 | 2.0 | 96.0 |
| Cefepime† | 1.9 | 7.4 | 90.7 | 9.7 | 3.2 | 87.1 |
| Meropenem | 100.0 | 0.0 | 0.0 | 98.0 | 2.0 | 0.0 |
| Gentamicin | 48.5 | - | 51.5 | 48.0 | - | 52.0 |
| Ciprofloxacin | 8.8 | 11.8 | 79.4 | 22.0 | 4.0 | 74.0 |
| Trimethoprim-sulfamethoxazole | 27.9 | 0.0 | 72.1 | 26.0 | 0.0 | 74.0 |
| Nitrofurantoin‡ | 100.0 | - | 0.0 | 89.5 | - | 10.5 |
| Tigecycline† | 100.0 | - | 0.0 | 100.0 | - | 0.0 |

S, susceptible; I, susceptible, increased exposure; R, Resistant; i.v. intravenous; *Breakpoints for uncomplicated urinary tract infections; †available for blood culture isolates only (n=85); ‡available for urine culture isolates only (n=33)

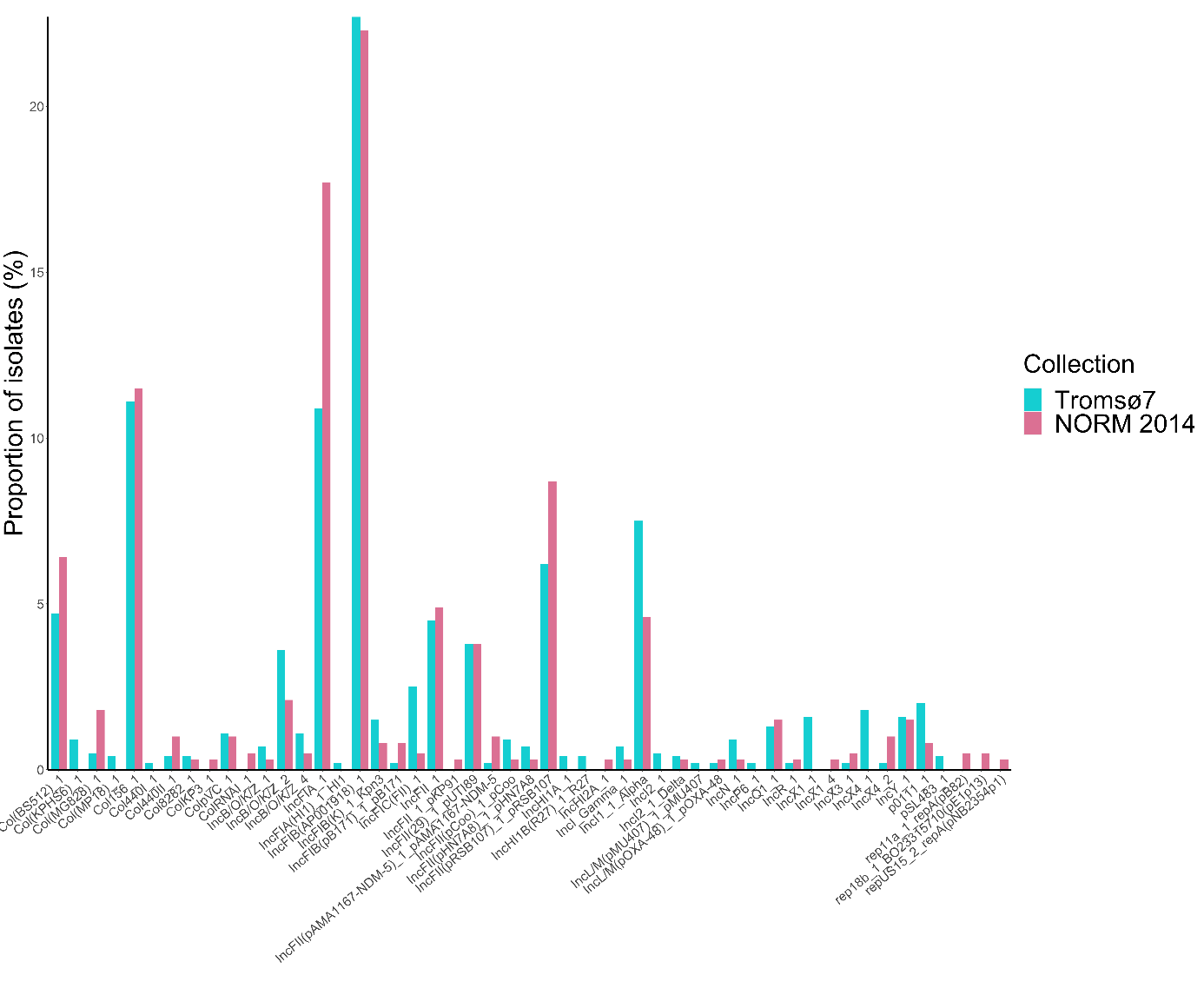
**Supplementary Figure 2.** Replicon type distribution among ESBL-*E. coli* carriage isolates from Tromsø7 (n=166) and clinical isolates from NORM 2014 (n=118).

**Supplementary Figure 3**.

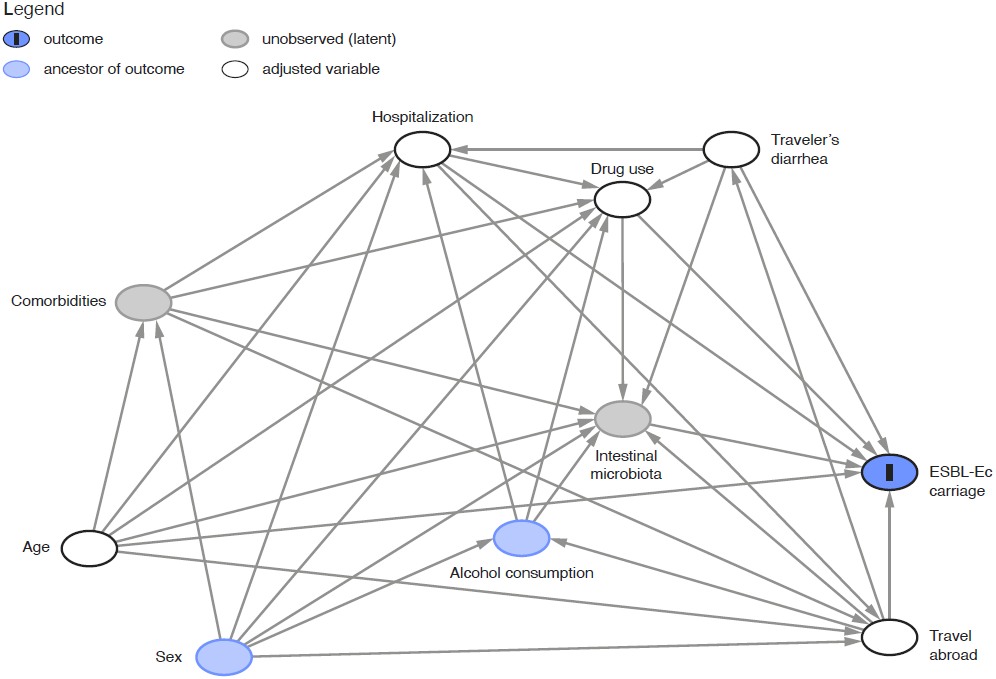

Direct acyclic graph (DAG) illustrating the causal relationship between ESBL-*E. coli* (ESBL-Ec) gastrointestinal carriage (outcome) and relevant covariates among 4,996 participants in Tromsø7. The variable ‘drug use’ includes antibiotic use past 14 days and acid suppressive medication past four weeks.

**Method:** To set up the most plausible causal relationship between covariates and the outcome, we first searched the literature for relevant factors associated with ESBL-*E. coli* gastrointestinal carriage. Thereafter, the DAG guided the selection of the multivariable logistic regression model, which was adjusted for the minimal sufficient adjustment set comprising age, drug use, hospitalization, travel abroad and traveler`s diarrhea (i.e. the variables constituting a confounding pathway). Controlling for the minimal sufficient adjustment set warrants that confounding paths are blocked in order to minimize bias of the causal relationship. The model was not adjusted for sex due to no statistically significant sex difference in prevalence of ESBL-*E. coli* carriage and because it does not constitute biasing paths after adjustment. For the variables sex and alcohol consumption, a direct effect on ESBL-*E. coli* carriage is not described or investigated, however they are known to affect the microbiota composition. Hence, intestinal microbiota was included as an unobserved mediator of the causal relationship.

**Supplementary Figure 4**.

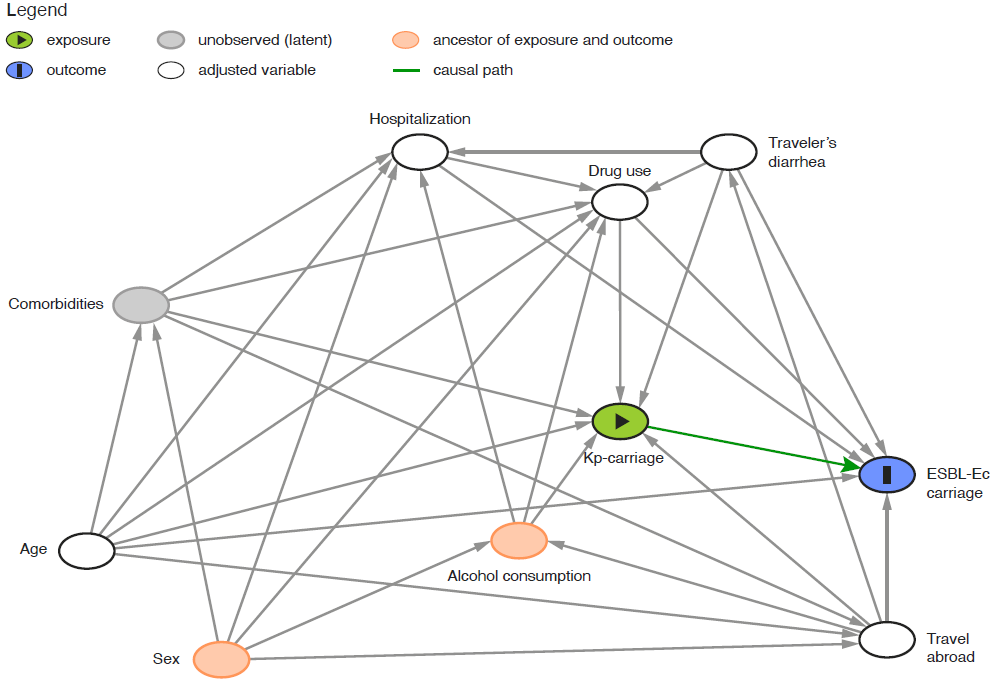

Direct acyclic graph (DAG) illustrating the causal relationship between *K. pneumoniae* species complex (Kp) gastrointestinal carriage (exposure), ESBL-*E. coli* (ESBL-Ec) gastrointestinal carriage (outcome) and relevant covariates in Tromsø7. The variable ‘drug use’ includes antibiotic use past 14 days and acid suppressive medication past four weeks.

**Method:** *K. pneumoniae* is a common cause of healthcare associated infections often combined with antimicrobial resistance. In the literature and our previous study^1^, we identified several factors associated with *K. pneumoniae* gastrointestinal carriage, which are overlapping with those associated with ESBL-*E. coli* carriage. To capture the relevance of *K. pneumoniae* carriage as an exposure, we included this variable into the causal relationship with ESBL-*E. coli* carriage. The DAG was used for the selection of variables for the multivariable logistic regression model, to conceptualize confounding and to identify the minimal sufficient adjustment set. Although sex and alcohol consumption represent ancestors of exposure and outcome, the adjustment for age, drug use, hospitalization, travel abroad and traveler`s diarrhea controls for relevant confounders and blocks biasing paths. The absence of red arrows in the DAG implies that there are no open unadjusted confounding pathways.
